# Prognosis of Acute Ischaemic Stroke in Cancer Patients: A National Inpatient Sample Study

**DOI:** 10.1101/2021.03.09.21253211

**Authors:** Tiberiu A Pana, Mohamed O Mohamed, Mamas A Mamas, Phyo K Myint

## Abstract

**Background and Purpose:** Whilst cancer is a risk factor for acute ischaemic stroke (AIS), its impact on AIS prognosis between metastatic and non-metastatic (MC and NMC) disease is poorly understood. Furthermore, the receipt of intravenous thrombolysis (IVT) and endovascular thrombectomy (ET) and their outcomes is poorly researched.

**Methods:** AIS admissions from the National Inpatient Sample (NIS) were included (October 2015-December 2017). Multivariable logistic regressions adjusting for a wide range of confounders analysed the relationship between NMC and MC and AIS in-hospital outcomes (mortality, prolonged hospitalisation >4 days and routine home discharge). Interaction terms with IVT and ET were also computed to explore their impact amongst cancer patients.

**Results:** 221,249 records representative of 1,106,045 admissions were included. There were 38,855 (3.51%) patients with co-morbid cancers: NMC=53.78% and MC=46.22%. NMC was associated with 23% increased odds of in-hospital mortality (odds ratio (95% confidence interval) = 1.23 (1.07-1.42)), which was mainly driven by pancreatic and respiratory cancers. This association was entirely offset by both IVT and ET. MC was associated with 2-fold increased odds of in-hospital mortality (2.16 (1.90-2.45)), which was mainly driven by respiratory, pancreatic and colorectal cancers. This association was only offset by ET. Both NMC and MC were significantly associated with prolonged hospitalisation and decreased odds of routine discharge.

**Conclusions:** Cancer patients are at higher odds of acute adverse outcomes after AIS and warrant robust primary prevention. IVT and ET improve these outcomes and should thus be offered routinely unless otherwise contraindicated in this group of stroke patients.

## INTRODUCTION

Malignancy is associated with increased risk of acute ischaemic stroke (AIS)^1^. This is mediated through a variety of mechanisms, including hypercoagulability^2^ and shared risk factors^1^. Cancer is therefore important to consider in AIS, as it may not only be more prevalent amongst AIS patients^3^, but also be associated with adverse outcomes^4,5^. Despite that fact that cancer is a heterogenous disease, the distribution of organ-specific primary cancer types amongst AIS patients and the magnitude of the association between each cancer type and adverse AIS outcomes remain largely unknown.

Revascularisation therapies (intravenous thrombolysis – IVT and endovascular thrombectomy - ET) significantly improve AIS outcomes^6,7^ and are thus recommended routinely in eligible patients^8^. Nevertheless, co-morbid cancer may hinder their use given that this population is more likely to exhibit contraindications to IVT or ET^1^. Current guidelines recommend IVT in AIS patients with systemic malignancy provided they have a life expectancy >6 months and no contraindications^8^. These recommendations are based on limited evidence, as landmark randomised controlled trials studying IVT^9–12^ or ET^7^ have excluded cancer patients. Previous smaller scale observational studies of AIS patients with cancer have found no increased haemorrhagic complications or mortality associated with IVT^13–15^. Nevertheless, guidelines currently provide no specific recommendations regarding ET for this population^8^, with the efficacy and safety of ET amongst AIS patients with cancer being largely unclear. While several small retrospective studies have found no association between cancer and adverse outcomes in AIS patients undergoing ET^16,17^, others have identified significantly higher mortality in cancer patients^18^.

This drives the need for a comprehensive description of the association between cancer and AIS outcomes in contemporary clinical practice and whether revascularisation therapies have an effect on these outcomes. In this study of a representative sample of AIS admissions in the United States (US) between 2015-2017, we sought examine the prevalence of comorbid cancer among patients with AIS and its association with in-hospital outcomes, also stratifying by metastatic disease. We also aimed to examine the effect of IVT/ET on outcomes in cancer patients through the use of interaction terms.

## METHODS

This study was conducted in accordance with the principles of the Declaration of Helsinki (1975) and later amendments. The data that support the findings of this study are available from the corresponding author upon reasonable request.

### Data source and inclusion criteria

The National Inpatient Sample (NIS) is a publicly available database containing >7 million annual hospital admission records. NIS contains admission records representing a 20% stratified sample of all community hospital admissions in the United States. Using the provided sampling weights, the NIS data can be used to provide national estimates for the sampling population, representative of ∼95% of the US population^19,20^. Prior to undertaking this project, all authors completed the Healthcare Cost and Utilisation Project (HCUP) Data Use Agreement Training Tool. All authors also read and signed the Data Use Agreement for Nationwide Databases. As the NIS is publicly available and contains no patient identifiable information, no ethical approval was needed. Using data files containing annual admissions between 2015-2017, all records with a primary diagnosis of ischaemic stroke (*International Classification of Disease* – *tenth edition* (ICD-10) codes I63.0-I63.9) were extracted. Only cases admitted between October 2015-December 2017 were included due to a change in co-morbidity coding (ICD-9 to ICD-10) occurring after September 2015^20^. Elective admissions and those with missing data on key variables were excluded.

### Statistical Analysis

All analyses were performed using Stata 15.1SE, Stata Statistical Software. A 5% threshold of statistical significance was utilised for all analyses (*P <* 0.05). Analyses were performed following HCUP guidelines^21^, utilising the provided discharge weights as probability weights and survey data analysis techniques stratifying by NIS stratum and year of admission^22^ in order to account for patient clustering within hospitals and produce US-wide estimates^23^.

#### Outcomes

The following outcomes were analysed: (1) in-hospital mortality, (2) prolonged hospital stay in excess of 4 days and (3) routine discharge from hospital. Vital status upon hospital discharge (dead/alive) and the length of stay (LoS) in hospital are provided as standard variables in the NIS^24,25^. Prolonged hospitalisation was defined as LoS >4 days, according to expert clinical opinion and previous studies assessing ischaemic stroke outcomes amongst patients admitted to hospital in the United States^26^. A dichotomous variable indicating patients hospitalised for >4 days was subsequently used as an outcome for LoS analyses. Discharge status was coded using the provided discharge destination^27^. All records of patients who were discharged against medical advice and those discharged to an unknown destination were excluded from the analyses prior to weighting (n=2187 (0.99 %)), allowing estimates for this particular outcome to be provided for 1,095,110 (99.01%) of AIS patients. Discharge destination was then dichotomised into routine discharges and other discharges (‘home health care’, ‘short-term hospital’, ‘other facilities including intermediate care and skilled nursing home’ and ‘died in hospital’). The ‘other discharges’ category was subsequently used as a reference category in all analyses evaluating discharge destination.

#### Exposures and confounders

Co-morbid cancer (non-metastatic and metastatic) as well as the organ-specific types were the exposures of interest. All models were adjusted for the following confounders: age, sex, ethnicity, Elixhauser co-morbidities (congestive heart failure, valvular disease, pulmonary circulatory disease, peripheral vascular disease, paralysis, other neurological disorders, chronic pulmonary disease, diabetes, hypothyroidism, renal failure, liver disease, peptic ulcer disease, acquired immune deficiency syndrome, rheumatoid arthritis, coagulopathy, obesity, weight loss, fluid and electrolyte disorders, anaemia, alcohol abuse, drug abuse, psychosis, depression and hypertension), previous history of cancer, haematological malignancies, other co-morbidities (dyslipidaemia, smoking, Parkinson disease, coronary heart disease, all-cause bleeding, pulmonary embolism, deep venous thrombosis, atrial fibrillation, arrhythmias other than atrial fibrillation, pneumonia (incl. aspiration), shock, previous cerebrovascular disease), hospital bedsize, location & teaching status and revascularisation therapy (thrombolysis, thrombectomy). Adjusting co-variates were selected based on clinical judgement and previous literature^5,14,15,28^.

Co-morbid non-metastatic and metastatic cancer diagnoses were identified using the Elixhauser co-morbidities^29^: solid tumour without metastases and metastatic cancer, respectively. Specific cancer types were identified using the Clinical Classification Software Refined (CCSR) codes (Supplemental Table I - please see https://www.ahajournals.org/journal/str)^30^. Previous history of cancer was identified using ICD10 codes Z85.x and Z86.0x. Elixhauser co-morbidities were determined using the HCUP Elixhauser co-morbidity software version 2020.1^29^. Co-morbid conditions other than the Elixhauser co-morbidities were identified using ICD-10 codes (Supplemental Table II - please see https://www.ahajournals.org/journal/str).

#### Descriptive Statistics

Patient characteristics were compared between AIS patients without cancer, those with non-metastatic cancer and those with metastatic disease. One-way analysis of variance and Pearson’s χ^2^ test were employed to compare patient characteristics for continuous and categorical variables, respectively.The distribution of each primary cancer type amongst the included sample as well as the proportion of metastatic disease amongst each cancer type were determined. Patient characteristics were then also compared between AIS patients with the 5 most common types of primary cancers previously identified. No between-group tests of statistical significance were performed for these comparisons as the groups were not mutually exclusive.

#### Association between prevalent cancer and odds of receiving revascularisation therapy

Multivariable logistic regressions were employed to analyse the relationship between co-morbid non-metastatic and metastatic cancer and the odds of receiving IVT and ET in hospital. All models were adjusted for the covariates listed above, with the exception of IVT or ET when this variable was used as the outcome.

#### Association between non-metastatic and metastatic cancer and in-hospital outcomes

Multivariable logistic regressions were employed to analyse the relationship between co-morbid non-metastatic and metastatic cancer and in-hospital outcomes. Separate models containing interaction terms with IVT and ET were also computed to determine whether these relationships were modified by revascularisation therapies. All models were adjusted for the covariates listed above.

#### Association between the five most common primary cancer types and in-hospital outcomes

Multivariable logistic regressions were employed to analyse the relationship between the five most common primary cancer types and in-hospital outcomes, stratifying each cancer type by the presence of metastases were also computed. All five cancer types were simultaneously introduced in the same model. All models were adjusted for the covariates listed above as well as other co-morbid cancer types.

## RESULTS

Figure 1 details the study population. Out of 230,177 records extracted with a primary diagnosis of ischaemic stroke between October 2015-December 2017, a total of 8708 elective admission records as well as 220 records with missing data were excluded, yielding a total of 221,249 included records. After the application of sampling weights and the exclusion of strata with single sampling units, the included records were used to provide estimates for the population from which they were sampled: 1,106,045 patients admitted with a primary diagnosis of AIS.

**Figure 1.**
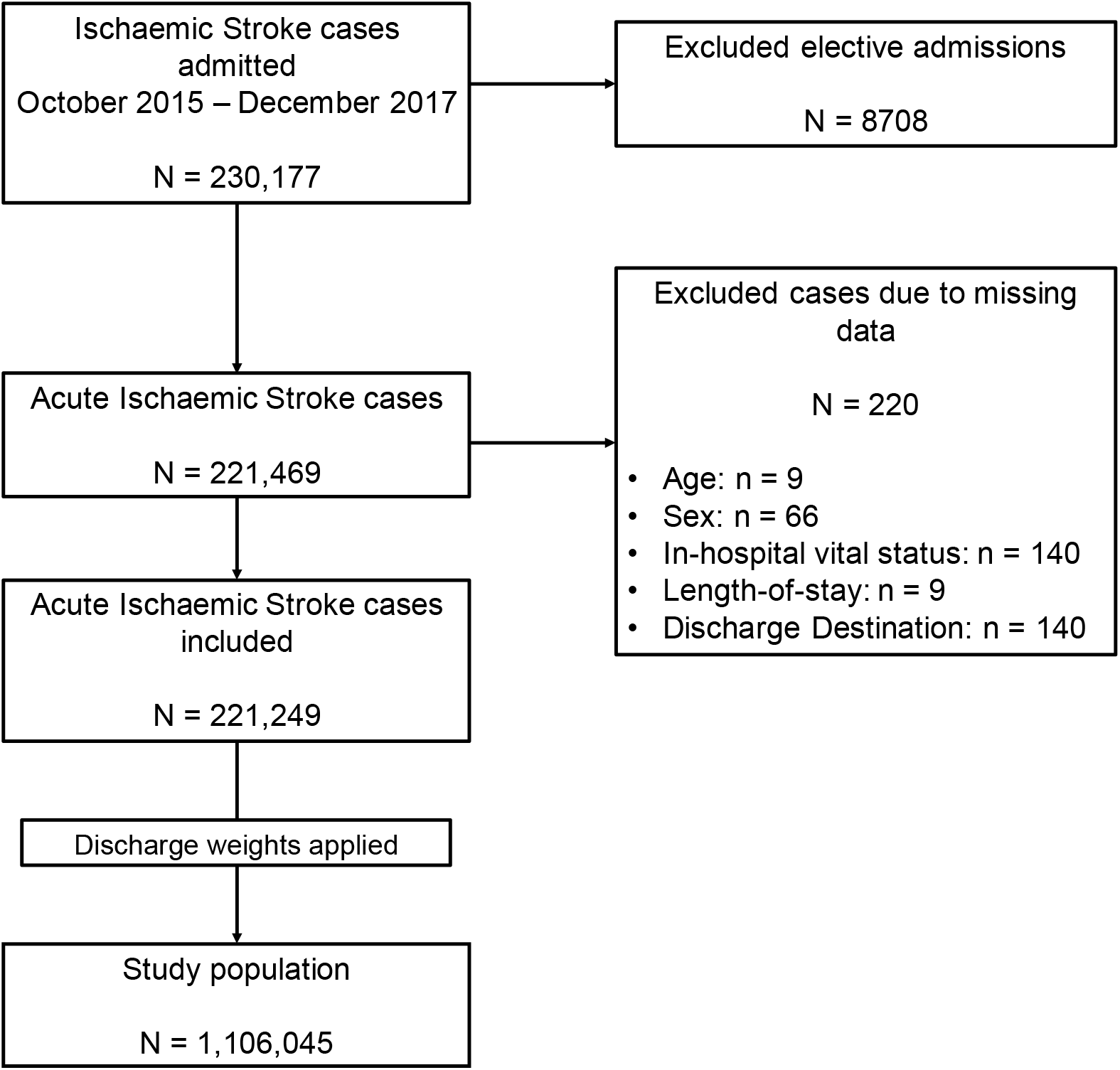
Patient Population Flowchart.

### Descriptive Statistics

Figure 2 details the distribution of primary cancer types amongst the 38,855 AIS patients with co-morbid cancer, representing 3.51% of the entire included sample. The five most common types were: respiratory (9490 (24.42%)), prostate (4960 (12.77%)), breast (3375 (8.69%)), pancreatic (2640 (6.79%)) and colorectal cancers (2490 (6.41%)). There were 4750 (50.05%) metastatic respiratory cancer patients, 1250 (25.20%) metastatic prostate cancer patients, 1110 (32.89%) metastatic breast cancer patients, 1875 (71.02%) metastatic pancreatic cancer patients and 985 (39.56%) metastatic colorectal cancer patients.

**Figure 2.**
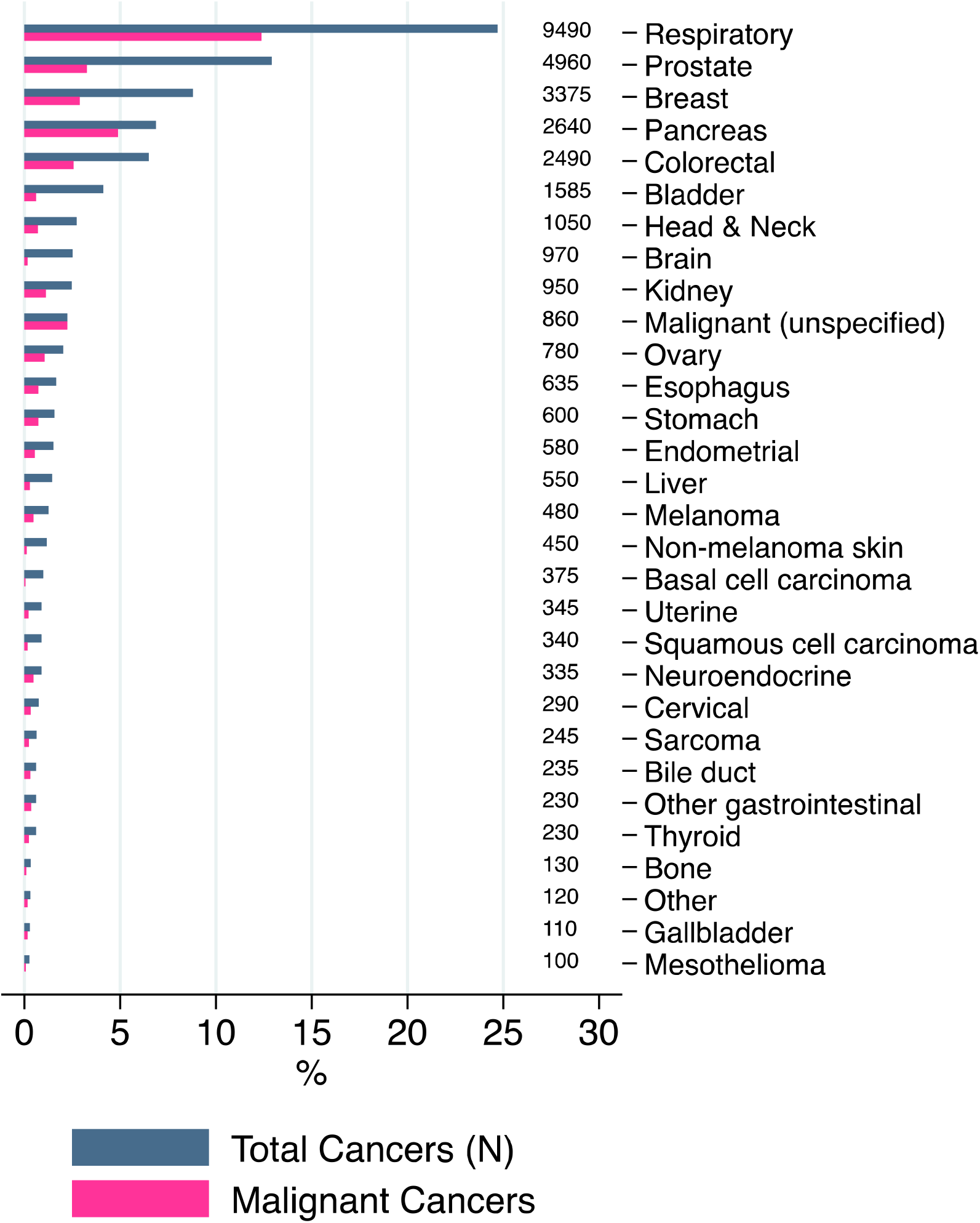
Distribution of each primary cancer type amongst the included sample of acute ischaemic stroke patients, representative of 1,106,045 patients.

Table 1 and Supplemental Table III (please see https://www.ahajournals.org/journal/str) detail the characteristics of the included patient population, representative of 1,106,045 AIS patients. The median (interquartile range) age was 72 (61-82) years and 50.41% were female. The median (interquartile range) length of stay was 3 (2-6) days. There were 20,895 (1.89%) patients with non-metastatic cancer and 17,960 (1.62%) with metastatic cancer. Median age ranged between 70 and 75 years, highest among those with non-metastatic cancer and lowest amongst those with metastatic cancer. The highest proportion of females was recorded amongst patients with metastatic cancer (51.03%), followed by those without active cancer (50.47%) and those with non-metastatic cancer (46.88%). Compared to patients without cancer, those with cancer had higher rates of prevalent chronic pulmonary disease, liver disease, coagulopathy and anaemia, but lower rates of congestive heart disease and diabetes. Compared to patients without cancer, those with cancer had higher rates of in-hospital mortality, prolonged hospitalisation and lower rates of routine discharge.

**Table 1.**
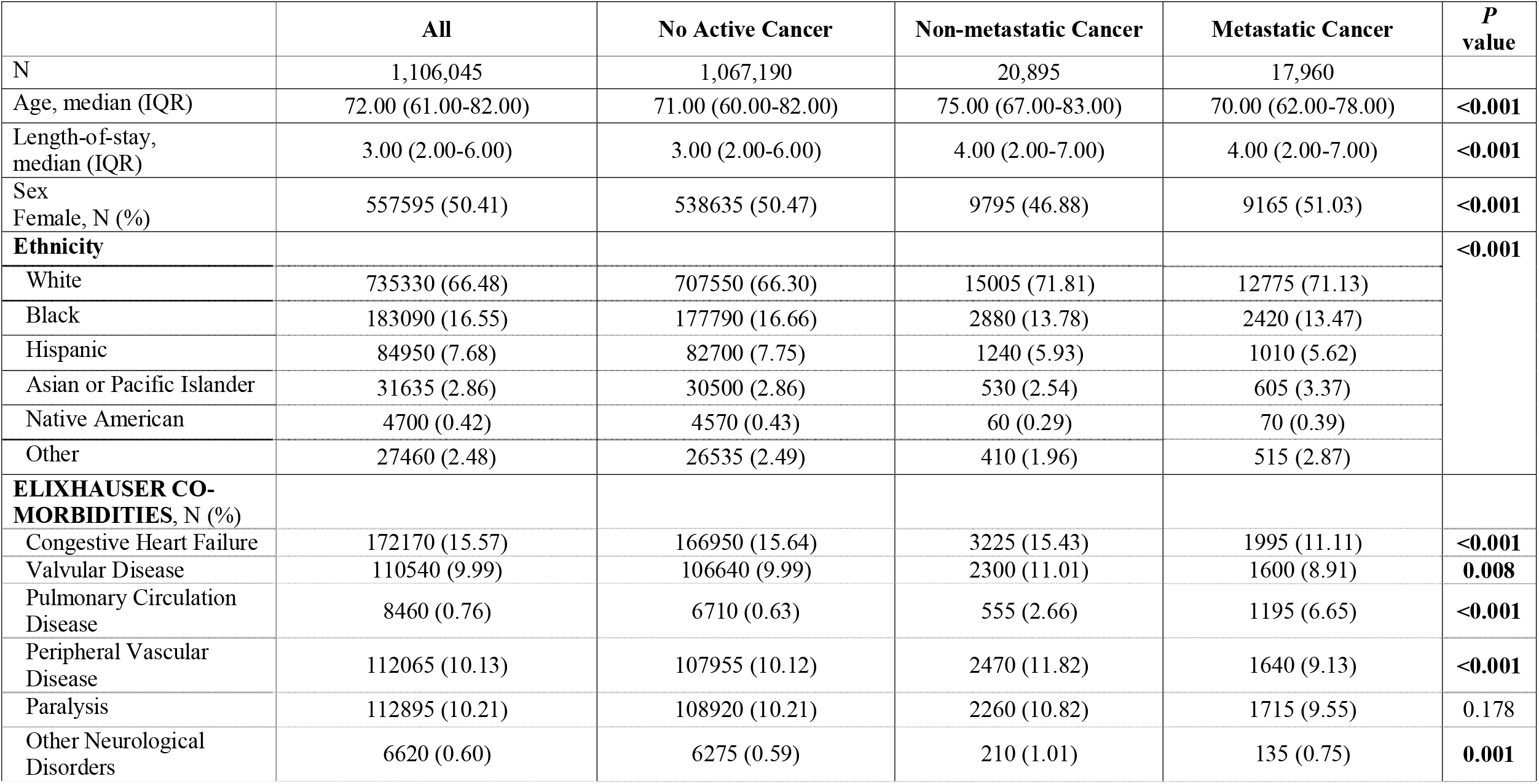
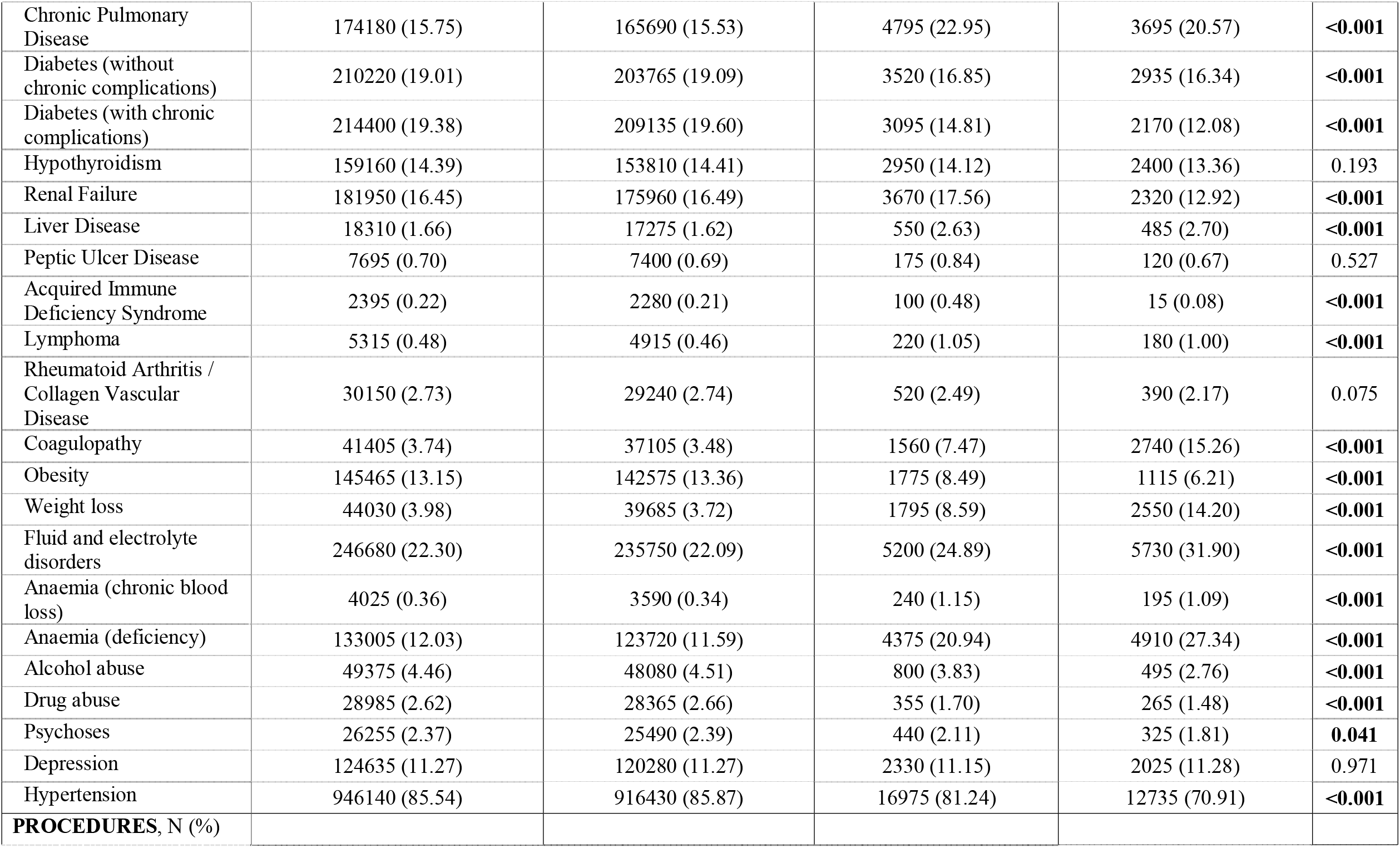
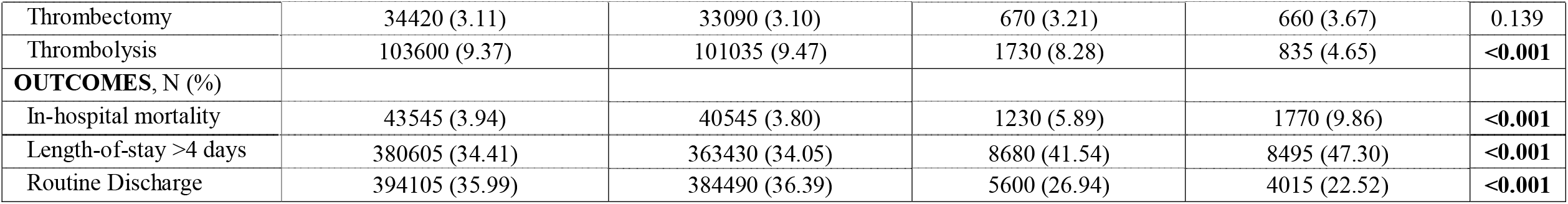
Patient characteristics on admission, stratified by either co-existent non-metastatic or metastatic cancer. Further descriptive statistics are detailed in Supplemental Table III (please see https://www.ahajournals.org/journal/str).

Supplemental Table IV (please see https://www.ahajournals.org/journal/str) details the characteristics of the included patients with the five most common co-morbid cancer types. Patients with prostate cancer were oldest, median (interquartile range) – 79 (71-85) years, followed by those with breast cancer – 74 (67-83) years, colorectal cancer – 74 (65-82) years, respiratory cancers – 71 (63-79) years and pancreatic cancer 71 (63-77) years. Patients with pancreatic cancer had the highest rate of in-hospital mortality (10.61%), followed by respiratory cancers (10.17%), colorectal cancer (6.22%), breast cancer (5.63%) and prostate cancer (3.73%).

### Association between prevalent cancer and odds of receiving revascularisation therapy

Supplemental Table V (please see https://www.ahajournals.org/journal/str) details the results of the multivariable logistic regressions evaluating the associations between non-metastatic and metastatic cancer and the odds of receiving thrombolysis or thrombectomy. Compared to patients without cancer, those with non-metastatic cancer had lower odds of receiving both IVT (odds ratio (95% confidence interval) – 0.77 (0.66-0.90)) and ET (0.80 (0.63-0.9994)). Compared to patients without cancer, those with metastatic cancer had lower odds of receiving IVT (0.39 (0.32-0.47)) but not ET (0.87 (0.69-1.09)).

### Association between non-metastatic and metastatic cancer and in-hospital outcomes

Figure 3 details the results of the multivariable logistic regressions assessing the associations between non-metastatic and metastatic cancer and in-hospital outcomes. Non-metastatic cancer was associated with a 23% increase in the odds of in-hospital mortality (odds ratio (95% confidence interval) – 1.23 (1.07-1.42)). This association exhibited significant interactions with IVT and ET: non-metastatic cancer was not associated with increased in-hospital mortality amongst AIS patients undergoing either IVT or ET. Metastatic cancer was associated with a 2-fold increase in the odds of in-hospital mortality: 2.16 (1.90-2.45). This association also exhibited a significant interaction with ET, but not IVT: metastatic cancer was not associated with increased in-hospital mortality amongst AIS patients undergoing ET. Non-metastatic and metastatic cancers were also associated with increased odds of prolonged hospitalisation and decreased odds of routine discharge.

**Figure 3.**
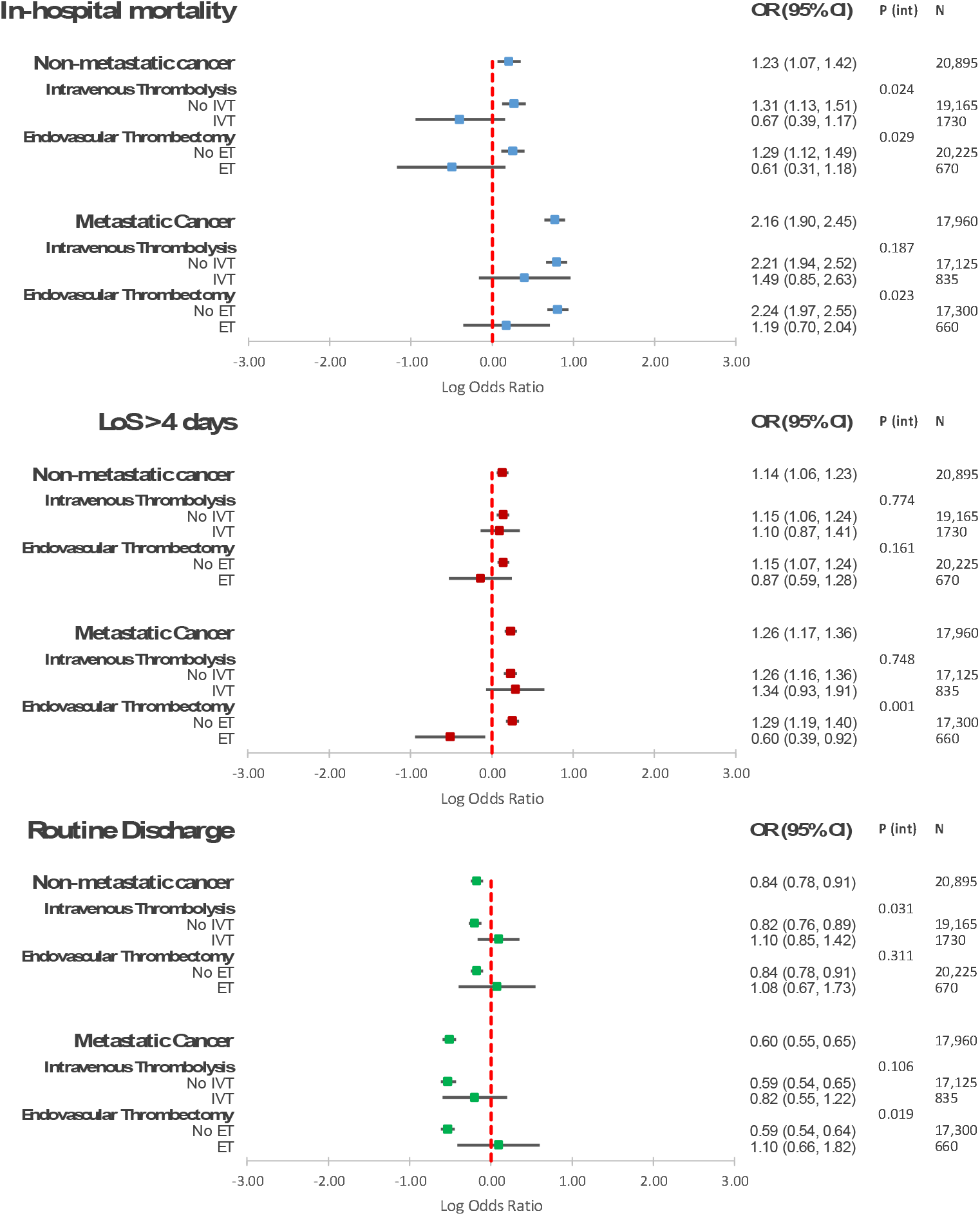
Results of multivariable logistic regressions assessing the association between co-morbid cancer (non-metastatic and metastatic) and acute ischaemic stroke in-hospital outcomes as well as the interaction with revascularisation therapies. OR – odds ratio; CI – confidence interval; IVT – intravenous thrombolysis; ET – endovascular thrombectomy

### Association between the five most common primary cancer types and in-hospital outcomes

Figure 4 details the results of the multivariable logistic regression assessing the association between the five most common primary cancer types and in-hospital outcomes, stratifying by metastatic disease status. Respiratory cancers (both non-metastatic – 1.88 (1.48-2.40) and metastatic – 2.40 (1.90-3.02)), pancreatic cancers (both non-metastatic (1.96 (1.04-3.71)) and metastatic – 2.33 (1.61-3.37)) and metastatic colorectal cancer (2.08 (1.21-3.58)) were associated with significantly increased in-hospital mortality. There were no associations between metastatic prostate cancer, breast cancer (both non-metastatic and metastatic) and non-metastatic colorectal cancer and in-hospital mortality. Non-metastatic prostate cancer was associated with decreased odds of in-hospital mortality (0.62 (0.40-0.96)). Respiratory cancers, metastatic pancreatic and metastatic colorectal cancer were associated with increased odds of prolonged hospitalisation. Respiratory cancers, metastatic prostate cancer, pancreatic cancer (both non-metastatic and metastatic) and metastatic colorectal cancer were associated with decreased odds of routine discharge.

**Figure 4.**
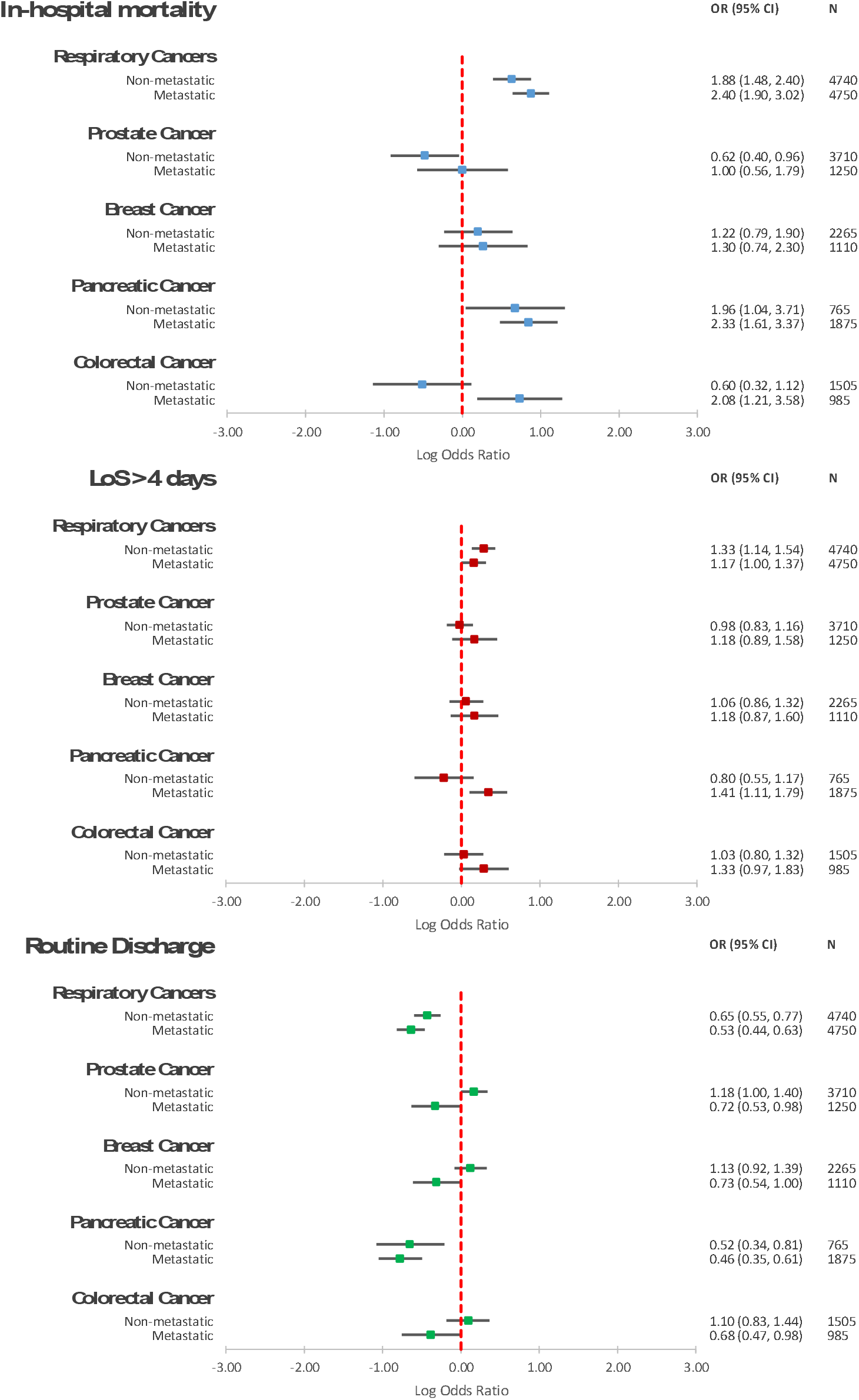
Results of multivariable logistic regressions assessing the association between the 5 most commonly occurring cancer types in the included cohort and acute ischaemic stroke in-hospital mortality as well as the interaction with revascularisation therapies. OR – odds ratio; CI – confidence interval; IVT – intravenous thrombolysis; ET – endovascular thrombectomy

## DISCUSSION

In this study including a sample representative of 1,106,045 acute ischemic stroke patients admitted between 2015-2017, we have determined the distribution of prevalent cancers as well as their association with in-hospital outcomes. We have also determined how these associations are influenced by the use of revascularisation therapies.

The five most common cancer types were: respiratory (24.42%), prostate (12.77%), breast (8.69%), pancreatic (6.79%) and colorectal cancers (6.41%). We report significant differences in stroke treatment according to cancer diagnosis, with patients with non-metastatic cancer at 23% lower odds of receiving IVT and 20% lower odds of receiving ET. The disparities in treatments were even greater in patients with metastatic cancer who were at 61% lower odds of receiving IVT compared to patients without cancer. Patients with cancer were at increased risk of adverse AIS outcomes. Non-metastatic cancer was associated with 23% increased odds of in-hospital mortality, which was mainly driven by pancreatic (96% increased odds) and respiratory (88% increased odds) cancers. This association was entirely offset by both IVT and ET. Metastatic cancer was associated with 2-fold increased odds of in-hospital mortality, which was mainly driven by respiratory (2.43-fold increase), pancreatic (2.37-fold increase) and colorectal (2.11-fold) cancers. This association was only offset by ET. Non-metastatic and metastatic cancers were also associated with increased odds of prolonged hospitalisation and decreased odds of routine discharge.

The association between co-morbid cancer and AIS outcomes has been previously evaluated on smaller, single-centre cohorts^4,31,32^. A previous investigation including ∼5000 AIS patients out of whom 1.46% had co-morbid cancer found a 3.7-fold increase in the odds of in-hospital mortality^4^. Another study including 468 AIS patients with co-morbid cancer found that metastatic disease was independently associated with a 4.5-fold increase in the risk of 6-month mortality compared to non-metastatic cancers^32^. Furthermore, gastric and pancreatic cancers were associated with increased mortality risk compared to other cancer types^32^. Our results complement these previous findings by providing a comprehensive description of the association between these disease entities based on a large, national real-world sample of AIS patients and highlight disparities in provision of evidence-based therapies.

Furthermore, our results provide additional insight into these relationships by exploring differences based on the presence of metastases and between different primary types. We found 23% increased odds of in-hospital mortality associated with non-metastatic cancer, which may be attributed to a higher proportion of cryptogenic strokes^4^ with worse prognosis^33^ and increased hypercoagulability leading to complications such as venous thromboembolism, recurrent stroke ^34^ and a greater risk of haemorrhagic transformation. Another important contributor to the increased risk of adverse outcomes was significant differences in the receipt of IVT and ET, with patients with cancer consistently less likely to receive revascularisation therapies.

Previous large clinical trials assessing the use of IVT for AIS revascularisation provide no specific information regarding AIS patients with cancer^9–12^. Nevertheless, several observational studies including AIS patients with co-morbid cancer have found no association between IVT and in-hospital mortality, major bleeding or functional outcomes^13,14,35–38^, while current guidelines recommend IVT in patients with systemic malignancy provided they have a life expectancy >6 months and no other contraindications^8^. We found that even after comprehensive adjustment, patients with co-morbid cancer were less likely to receive IVT, suggesting that the lower rates of IVT in this population may not be fully explained by a higher prevalence of contraindications. This may reflect the fact that treating clinicians may be hesitant to use IVT solely based on cancer status, even in those with non-metastatic disease.

Our work shows for the first time that IVT offset the increased in-hospital mortality and decreased odds of routine discharge associated with non-metastatic cancer, providing supportive evidence for IVT therapy in these patients. Nevertheless, the associations between metastatic cancer and adverse in-hospital outcomes were not offset by IVT, suggesting that AIS patients with metastatic cancer may not fully benefit from this therapy. This may relate to the fact that strokes in this patient group may also relate to the presence of cerebral metastases and not due to an in-situ thrombosis in the cerebral vessels, or that there was a greater likelihood of haemorrhagic transformation that offset any benefit with thrombolysis.

As with IVT, clinical trials assessing the use of ET in AIS provide no specific data regarding its use in cancer patients^7^. Nevertheless, a few small retrospective observational studies have assessed the use of ET in Asian AIS patients with cancer, reaching different conclusions^16,17,32^. While similar acute outcomes were found amongst AIS patients with and without co-morbid cancer undergoing ET in two studies^16,17^, a third study found significantly worse functional outcomes in cancer patients treated with ET^18^. Furthermore, it has been postulated that AIS patients with cancer may have different clot composition, which may hinder successful recanalisation in this population^16^.

Our analysis shows that patients with non-metastatic cancer were significantly less likely to receive ET, while those with metastatic disease were equally likely to receive ET compared to patients without cancer. These differences may reflect the fact that metastatic cancer patients may be more likely to present with cardioembolic strokes caused large artery occlusion, rendering them more likely candidates for ET. Nevertheless, in the absence of more granular stroke syndrome data, we could not assess this hypothesis and further research is thus warranted. Our study provides for the first time an analysis of the relationship between cancer and ET in AIS patients using a large, real-world and contemporary sample. ET offset the excess odds of in-hospital adverse outcomes associated both with non-metastatic and metastatic cancers, suggesting that ET may be a successful strategy in eligible patients with cancer, especially in those with metastases who may not fully benefit from IVT.

Our study has several strengths, such as including a large sample representative of >1 million AIS patients admitted between late 2015-2017 across the United States as well as having adjusted for a wide range of important confounders. Our results thus reflect contemporary stroke management, including the more widespread adoption of ET and thus allow the generalisation of clinical implications to patients with similar characteristics. Our findings show that both non-metastatic and metastatic disease are associated with significant increases in in-hospital mortality, prolonged hospitalisation and decreased odds of routine home discharge. This highlights that cancer patients warrant thorough primary prevention, since they are not only more likely to suffer an incident stroke, but also at higher odds of acute adverse stroke outcomes. Given our large sample size, our study is able to provide more granular information regarding individual associations between each primary cancer type and adverse acute outcomes.

Our study highlights inequalities in the receipt of evidence-based reperfusion therapies in cancer patients Such differences have also been described in other cardiovascular conditions such as myocardial infarction^28^. We report that cancer patients offered treatment with IVT or ET may derive a benefit and that IVT may offset the non-metastatic cancer-associated excess odds of adverse outcomes. Furthermore, ET offsets the excess odds associated with both non-metastatic and metastatic disease. Along with previous findings, our study also suggests that co-morbid cancer should not represent a contraindication to AIS revascularisation therapies in itself.

Naturally, our study also has limitations. Having used administrative data, we defined AIS using ICD-10 codes and thus lacked more detailed information regarding stroke severity, or classification. Nevertheless, all analyses were adjusted for a wide range of confounders including some important predictors of severe or cardioembolic stroke, such as atrial fibrillation and heart failure^39,40^, which may have partly accounted for stroke severity or classification. Furthermore, we also lacked more information regarding cancer staging except for metastases. We were thus unable to further stratify our analyses by cancer stage. Our database also did not capture treatments such as antithrombotic therapy, which may contribute to the differences in outcomes. Finally, our study only assessed in-hospital outcomes and further research including is also required to characterise the long-term stroke outcomes after hospital discharge associated with co-morbid cancer as well as their interaction with revascularisation strategies.

In conclusion, in this study of a sample representative of 1.1 million AIS admissions across the United States between 2015-2017, we reported that patients with cancer represent one in thirty acute stroke admissions in the United States and are associated with an increased risk of mortality. We also report that even after adjustment for differences in comorbidity, patients with cancer are less likely to be offered revascularisation therapies. These disparities in care may contribute to some of the observed adverse outcomes associated with a cancer diagnosis. Nevertheless, IVT offset the non-metastatic cancer-associated excess odds of mortality, while ET offset both the non-metastatic and metastatic cancer-associated excess odds of mortality. Both non-metastatic and metastatic cancers were associated with increased odds of prolonged hospitalisation and decreased odds of routine discharge. IVT and ET are useful strategies to improve in-hospital outcomes in this population and should be offered routinely in cancer patients unless otherwise contraindicated.

## Data Availability

The data that support the findings of this study are available from the corresponding author upon reasonable request.

## ACKNOWLEDGEMENTS

We thank Dr Jesus Perdomo-Lampignano for his assistance with the figures. We also acknowledge the HCUP Data Partners (https://www.hcup-us.ahrq.gov/db/hcupdatapartners.jsp). TAP, PKM and MAM conceived the study. Data were analysed by TAP. TAP and PKM drafted the article, and all the authors contributed to writing the article. PKM is the guarantor.

## SUPPLEMENTAL MATERIALS

Supplemental Tables I-V.

### NON-STANDARD ABBREVIATIONS AND ACRONYMS

AIS: Acute Ischaemic Stroke
ET: Endovascular Thrombectomy
HCUP: Healthcare Cost and Utilisation Project
ICD-19: International Classification of Disease – tenth edition
IVT: Intravenous Thrombolysis
LoS: Length of stay
MC: Metastatic Cancer
NIS: National Inpatient Sample
NMC: Non-metastatic Cancer
US: United States;

## REFERENCES

1. Navi BB, Iadecola C. Ischemic stroke in cancer patients: A review of an underappreciated pathology. Ann. Neurol. 2018;83:873–883.

2. Navi BB, Reiner AS, Kamel H, Iadecola C, Elkind MS V, Panageas KS, DeAngelis LM. Association between incident cancer and subsequent stroke. Ann. Neurol. 2015;77:291–300.

3. Wilbers J, Sondag L, Mulder S, Siegerink B, van Dijk EJ. Cancer prevalence higher in stroke patients than in the general population: the Dutch String-of-Pearls Institute (PSI) Stroke study. Eur. J. Neurol. 2020;27:85–91.

4. Kneihsl M, Enzinger C, Wunsch G, Khalil M, Culea V, Urbanic-Purkart T, Payer F, Niederkorn K, Fazekas F, Gattringer T. Poor short-term outcome in patients with ischaemic stroke and active cancer. J. Neurol. 2016;263:150–156.

5. Corraini P, Szepligeti SK, Henderson VW, Ording AG, Horvath-Puho E, Sorensen HT. Comorbidity and the increased mortality after hospitalization for stroke: a population-based cohort study. J. Thromb. Haemost. 2018;16:242–252.

6. Wardlaw JM, Murray V, Berge E, del Zoppo GJ. Thrombolysis for acute ischaemic stroke. Cochrane database Syst. Rev. 2014;CD000213.

7. Goyal M, Menon BK, van Zwam WH, Dippel DWJ, Mitchell PJ, Demchuk AM, Davalos A, Majoie CBLM, van der Lugt A, de Miquel MA, et al. Endovascular thrombectomy after large-vessel ischaemic stroke: a meta-analysis of individual patient data from five randomised trials. Lancet (London, England). 2016;387:1723– 1731.

8. Powers WJ, Rabinstein AA, Ackerson T, Adeoye OM, Bambakidis NC, Becker K, Biller J, Brown M, Demaerschalk BM, Hoh B, et al. Guidelines for the Early Management of Patients With Acute Ischemic Stroke: 2019 Update to the 2018 Guidelines for the Early Management of Acute Ischemic Stroke: A Guideline for Healthcare Professionals From the American Heart Association/American Stroke. Stroke. 2019;50:e344–e418.

9. Hacke W, Kaste M, Bluhmki E, Brozman M, Davalos A, Guidetti D, Larrue V, Lees KR, Medeghri Z, Machnig T, et al. Thrombolysis with alteplase 3 to 4.5 hours after acute ischemic stroke. N. Engl. J. Med. 2008;359:1317–1329.

10. National Institute of Neurological Disorders and Stroke rt-PA Stroke Study Group. Tissue plasminogen activator for acute ischemic stroke. N. Engl. J. Med. 1995;333:1581–1587.

11. Sandercock P, Wardlaw JM, Lindley RI, Dennis M, Cohen G, Murray G, Innes K, Venables G, Czlonkowska A, Kobayashi A, et al. The benefits and harms of intravenous thrombolysis with recombinant tissue plasminogen activator within 6 h of acute ischaemic stroke (the third international stroke trial [IST-3]): a randomised controlled trial. Lancet (London, England). 2012;379:2352–2363.

12. Bluhmki E, Chamorro A, Davalos A, Machnig T, Sauce C, Wahlgren N, Wardlaw J, Hacke W. Stroke treatment with alteplase given 3.0-4.5 h after onset of acute ischaemic stroke (ECASS III): additional outcomes and subgroup analysis of a randomised controlled trial. Lancet. Neurol. 2009;8:1095–1102.

13. Inohara T, Liang L, Kosinski AS, Smith EE, Schwamm LH, Hernandez AF, Bhatt DL, Fonarow GC, Peterson ED, Xian Y. Thrombolytic therapy in older acute ischemic stroke patients with gastrointestinal malignancy or recent bleeding. Eur. stroke J. 2020;5:47–55.

14. Weeda ER, Bohm N. Association between comorbid cancer and outcomes among admissions for acute ischemic stroke receiving systemic thrombolysis. Int. J. Stroke. 2019;14:48–52.

15. Murthy SB, Karanth S, Shah S, Shastri A, Rao CPV, Bershad EM, Suarez JI. Thrombolysis for acute ischemic stroke in patients with cancer: a population study. Stroke. 2013;44:3573–3576.

16. Jung S, Jung C, Hyoung Kim J, Se Choi B, Jung Bae Y, Sunwoo L, Geol Woo H, Young Chang J, Joon Kim B, Han M-K, et al. Procedural and clinical outcomes of endovascular recanalization therapy in patients with cancer-related stroke. Interv. Neuroradiol. 2018;24:520–528.

17. Cho B-H, Yoon W, Kim J-T, Choi K-H, Kang K-W, Lee J-H, Cho K-H, Park M-S. Outcomes of endovascular treatment in acute ischemic stroke patients with current malignancy. Neurol. Sci. 2020;41:379–385.

18. Lee D, Lee DH, Suh DC, Kwon HS, Jeong D-E, Kim J-G, Lee J-S, Kim JS, Kang D-W, Jeon S-B, et al. Intra-arterial thrombectomy for acute ischaemic stroke patients with active cancer. J. Neurol. 2019;266:2286–2293.

19. Mohamed MO, Kirchhof P, Vidovich M, Savage M, Rashid M, Kwok CS, Thomas M, El Omar O, Al Ayoubi F, Fischman DL, et al. Effect of Concomitant Atrial Fibrillation on In-Hospital Outcomes of Non-ST-Elevation-Acute Coronary Syndrome-Related Hospitalizations in the United States. Am. J. Cardiol. 2019;124:465–475.

20. (HCUP) Healthcare Cost and Utilization Project. NIS Database Documentation [Internet]. Agency Healthc. Res. Qual. Rockville, MD. 2018 [cited 2020 Apr 28];Available from: https://www.hcup-us.ahrq.gov/db/nation/nis/nisdbdocumentation.jsp

21. (HCUP) Healthcare Cost and Utilization Project. Checklist for Working with the NIS [Internet]. Agency Healthc. Res. Qual. Rockville, MD. 2017 [cited 2020 Apr 28];Available from: www.hcup-us.ahrq.gov/db/nation/nis/nischecklist.jsp.

22. (HCUP) Healthcare Cost and Utilization Project. HCUP NIS Description of Data Elements [Internet]. Agency Healthc. Res. Qual. Rockville, MD. 2008 [cited 2020 Apr 28];Available from: www.hcup-us.ahrq.gov/db/vars/nis_stratum/nisnote.jsp.

23. Houchens R, Ross D, Elixhauser A. Final Report on Calculating National Inpatient Sample (NIS) Variances for Data Years 2012 and Later. 2015;

24. (HCUP) Healthcare Cost and Utilization Project. HCUP NIS Description of Data Elements [Internet]. Agency Healthc. Res. Qual. Rockville, MD. 2008 [cited 2020 Jun 28];Available from: https://www.hcup-us.ahrq.gov/db/vars/died/nisnote.jsp

25. (HCUP) Healtcare Cost and Utilization Project. HCUP NIS Description of Data Elements [Internet]. Agency Healthc. Res. Qual. Rockville, MD. 2008 [cited 2020 Jun 28];Available from: https://www.hcup-us.ahrq.gov/db/vars/los/nisnote.jsp

26. Myint PK, Sheng S, Xian Y, Matsouaka RA, Reeves MJ, Saver JL, Bhatt DL, Fonarow GC, Schwamm LH, Smith EE. Shock Index Predicts Patient-Related Clinical Outcomes in Stroke. J. Am. Heart Assoc. 2018;7:e007581.

27. (HCUP) Healthcare Cost and Utilization Project. HCUP NIS Description of Data Elements [Internet]. Agency Healthc. Res. Qual. Rockville, MD. 2008 [cited 2020 Apr 28];Available from: www.hcup-us.ahrq.gov/db/vars/dispuniform/nisnote.jsp.

28. Bharadwaj A, Potts J, Mohamed MO, Parwani P, Swamy P, Lopez-Mattei JC, Rashid M, Kwok CS, Fischman DL, Vassiliou VS, et al. Acute myocardial infarction treatments and outcomes in 6.5 million patients with a current or historical diagnosis of cancer in the USA. Eur. Heart J. 2020;41:2183–2193.

29. (HCUP) Healthcare Cost and Utilization Project. Tools Archive for Elixhauser Comorbidity Software Refined for ICD-10-CM [Internet]. Agency Healthc. Res. Qual. Rockville, MD. 2020 [cited 2021 Jan 30];Available from: https://www.hcup-us.ahrq.gov/toolssoftware/comorbidityicd10/comorbidity_icd10_archive.jsp

30. Healthcare Cost and Utilization Project (HCUP). Clinical Classifications Software Refined (CCSR). Agency Healthc. Res. Qual. Rockville, MD. 2020;

31. Cutting S, Wettengel M, Conners JJ, Ouyang B, Busl K. Three-Month Outcomes Are Poor in Stroke Patients with Cancer Despite Acute Stroke Treatment. J. Stroke Cerebrovasc. Dis. 2017;26:809–815.

32. Yoo J, Nam HS, Kim YD, Lee HS, Heo JH. Short-Term Outcome of Ischemic Stroke Patients With Systemic Malignancy. Stroke. 2019;50:507–511.

33. Navi BB, Singer S, Merkler AE, Cheng NT, Stone JB, Kamel H, Iadecola C, Elkind MS V, DeAngelis LM. Cryptogenic subtype predicts reduced survival among cancer patients with ischemic stroke. Stroke. 2014;45:2292–2297.

34. Navi BB, Singer S, Merkler AE, Cheng NT, Stone JB, Kamel H, Iadecola C, Elkind MS V, DeAngelis LM. Recurrent thromboembolic events after ischemic stroke in patients with cancer. Neurology. 2014;83:26–33.

35. Murthy SB, Biffi A, Falcone GJ, Sansing LH, Torres Lopez V, Navi BB, Roh DJ, Mandava P, Hanley DF, Ziai WC, et al. Antiplatelet Therapy After Spontaneous Intracerebral Hemorrhage and Functional Outcomes. Stroke. 2019;50:3057–3063.

36. Sobolewski P, Brola W, Szczuchniak W, Fudala M, Sobota A. Safety of intravenous thrombolysis for acute ischaemic stroke including concomitant neoplastic disease sufferers -experience from Poland. Int. J. Clin. Pract. 2015;69:666–673.

37. Masrur S, Abdullah AR, Smith EE, Hidalgo R, El-Ghandour A, Rordorf G, Schwamm LH. Risk of thrombolytic therapy for acute ischemic stroke in patients with current malignancy. J. Stroke Cerebrovasc. Dis. 2011;20:124–130.

38. Huang S, Lu X, Tang L V, Hu Y. Efficacy and safety of intravenous thrombolysis for acute ischemic stroke in cancer patients: a systemic review and meta-analysis. Am. J. Transl. Res. 2020;12:4795–4806.

39. Arboix A, Alió J. Cardioembolic stroke: clinical features, specific cardiac disorders and prognosis. Curr. Cardiol. Rev. 2010;6:150–161.

40. Pana TA, McLernon DJ, Mamas MA, Bettencourt-Silva JH, Metcalf AK, Potter JF, Myint PK. Individual and Combined Impact of Heart Failure and Atrial Fibrillation on Ischemic Stroke Outcomes. Stroke. 2019;50:1838–1845.

