## Supplementary Material for "Prognosis of Acute Ischaemic Stroke in Cancer Patients: A National Inpatient Sample Study"

### SUPPLEMENTAL MATERIAL

#### Supplemental Tables

**Supplemental Table I.** International Classification of Disease – tenth edition (ICD-10) and Clinical Classifications Software Refined (CCSR) codes used to extract admission co-morbidities and procedures.

| Co-morbidities | ICD-10 codes (Diagnosis) |
|---|---|
| Chronic Lung Disease | J43.x – J47.x; J60.x – J67.x; J68.4; J68.8; J68.9; J84.0x; J84.111 – J84.113; J84.115 – J84.17; J84x |
| Coronary Heart Disease | I20.x – I25.x |
| Deep Venous Thrombosis | I82.x |
| Pulmonary Embolism | I26.x |
| Pericarditis | I30.x – I32 |
| Infectious Endocarditis | I33.0; I33.9 |
| Atrial Fibrillation/Flutter | I48.x |
| Pneumonia | J12.x – J18.x |
| All-cause bleeding | D69.8; D69.9; G97.x; H11.3x; H31.3x; H35.6x; H43.1x; H92.2x; I31.2; I60.x—I62.x; I85.01; I85.11; K25.x-K29.x; K31.811; K62.5; K92.0-K92.2 I97.418; I97.42; I97.618-I97.621; J60.x-J62.x; N93.8; N93.9; N95.0; R04.x; R31.0; R31.9; R58; S06.4x – S06.6x |
| Shock | R57.x |
| Previous Cerebrovascular Disease | Z86.73 |
| Family History of Stroke | Z82.3 |
| Family History of Coronary Heart Disease | Z82.41; Z82.49 |
| Smoking | F17.2x |
| Dyslipidaemia | E78.0-E78.6 |
| **Procedures** | **ICD-10 codes (Procedural)** |
| Thrombolysis | 03CG3ZZ; 03CG4ZZ; 03CK3Z7; 03CK3ZZ; 03CK4ZZ; 03CL3Z7; 03CL3ZZ; 03CL4ZZ; 03CP3Z7; 03CP3ZZ; 03CP4ZZ; 03CQ3Z7; 03CQ3ZZ; 03CQ4ZZ |
| Thrombectomy | 03CG3ZZ; 03CG4ZZ; 03CK3Z7 |

ICD-10 - International Classification of Disease – tenth edition

**Supplemental Table II.** Clinical Classifications Software Refined (CCSR) codes used to classify cancer types.

| Cancer Type | CCSR code | CCSR description |
|---|---|---|
| Head and neck cancers | NEO001 | Head and neck cancers - eye |
| | NEO002 | Head and neck cancers - lip and oral cavity |
| | NEO003 | Head and neck cancers - throat |
| | NEO004 | Head and neck cancers - salivary gland |
| | NEO005 | Head and neck cancers - nasopharyngeal |
| | NEO006 | Head and neck cancers - hypopharyngeal |
| | NEO007 | Head and neck cancers - pharyngeal |
| | NEO008 | Head and neck cancers - laryngeal |
| | NEO009 | Head and neck cancers - tonsils |
| | NEO010 | Head and neck cancers - all other types |
| Cardiac cancers | NEO011 | Cardiac cancers |
| Gastrointestinal cancers - other | NEO012 | Gastrointestinal cancers - esophagus |
| | NEO013 | Gastrointestinal cancers - stomach |
| | NEO014 | Gastrointestinal cancers - small intestine |
| | NEO016 | Gastrointestinal cancers - anus |
| | NEO017 | Gastrointestinal cancers - liver |
| | NEO018 | Gastrointestinal cancers - bile duct |
| | NEO019 | Gastrointestinal cancers - gallbladder |
| | NEO020 | Gastrointestinal cancers - peritoneum |
| | NEO021 | Gastrointestinal cancers - all other types |
| Gastrointestinal cancers - colorectal | NEO015 | Gastrointestinal cancers - colorectal |
| Respiratory cancers | NEO022 | Respiratory cancers |
| Bone cancer | NEO023 | Bone cancer |
| Sarcoma | NEO024 | Sarcoma |
| Skin cancers - melanoma | NEO025 | Skin cancers - melanoma |
| Skin cancers - nonmelanoma | NEO026 | Skin cancers - basal cell carcinoma |
| | NEO027 | Skin cancers - squamous cell carcinoma |
| | NEO028 | Skin cancers - all other types |
| Breast cancer | NEO029 | Breast cancer - ductal carcinoma in situ (DCIS) |
| | NEO030 | Breast cancer - all other types |
| Female reproductive system cancers | NEO031 | Female reproductive system cancers - uterus |

| | | |
|---|---|---|
| | NEO032 | Female reproductive system cancers - cervix |
| | NEO033 | Female reproductive system cancers - ovary |
| | NEO034 | Female reproductive system cancers - fallopian tube |
| | NEO035 | Female reproductive system cancers - endometrium |
| | NEO036 | Female reproductive system cancers - vulva |
| | NEO037 | Female reproductive system cancers - vagina |
| | NEO038 | Female reproductive system cancers - all other types |
| Male reproductive system cancers | NEO039 | Male reproductive system cancers - prostate |
| | NEO040 | Male reproductive system cancers - testis |
| | NEO041 | Male reproductive system cancers - penis |
| | NEO042 | Male reproductive system cancers - all other types |
| Urinary system cancers | NEO043 | Urinary system cancers - bladder |
| | NEO044 | Urinary system cancers - ureter and renal pelvis |
| | NEO045 | Urinary system cancers - kidney |
| | NEO046 | Urinary system cancers - urethra |
| | NEO047 | Urinary system cancers - all other types |
| Nervous system cancers | NEO048 | Nervous system cancers - brain |
| | NEO049 | Nervous system cancers - all other types |
| Endocrine system cancers | NEO050 | Endocrine system cancers - thyroid |
| | NEO051 | Endocrine system cancers - pancreas |
| | NEO052 | Endocrine system cancers - thymus |
| | NEO053 | Endocrine system cancers - adrenocortical |
| | NEO054 | Endocrine system cancers - parathyroid |
| | NEO055 | Endocrine system cancers - pituitary gland |
| | NEO056 | Endocrine system cancers - all other types |
| Lymphoma | NEO057 | Hodgkin lymphoma |
| | NEO058 | Non-Hodgkin lymphoma |
| Leukemia | NEO059 | Leukemia - acute lymphoblastic leukemia (ALL) |

| | | |
|---|---|---|
| | NEO060 | Leukemia - acute myeloid leukemia (AML) |
| | NEO061 | Leukemia - chronic lymphocytic leukemia (CLL) |
| | NEO062 | Leukemia - chronic myeloid leukemia (CML) |
| | NEO063 | Leukemia - hairy cell |
| | NEO064 | Leukemia - all other types |
| Multiple myeloma | NEO065 | Multiple myeloma |
| Malignant neuroendocrine tumors | NEO066 | Malignant neuroendocrine tumors |
| Mesothelioma | NEO067 | Mesothelioma |
| Myelodysplastic syndrome (MDS) | NEO068 | Myelodysplastic syndrome (MDS) |
| Cancer of other sites | NEO069 | Cancer of other sites |
| Secondary malignancies | NEO070 | Secondary malignancies |
| Malignant neoplasm, unspecified | NEO071 | Malignant neoplasm, unspecified |
| Neoplasms of unspecified nature or uncertain behavior | NEO072 | Neoplasms of unspecified nature or uncertain behavior |
| Benign neoplasms | NEO073 | Benign neoplasms |
| Conditions due to neoplasm or the treatment of neoplasm | NEO074 | Conditions due to neoplasm or the treatment of neoplasm |

**Supplemental Table III**. Patient characteristics on admission, stratified by either co-existent non-metastatic or metastatic cancer.

| | All | No cancer | Non-metastatic Cancer | Metastatic Cancer | *P* value |
|---|---|---|---|---|---|
| N | 1106045 | 1067190 | 20895 | 17960 | |
| Age, median (IQR) | 72.00 (61.00-82.00) | 71.00 (60.00-82.00) | 75.00 (67.00-83.00) | 70.00 (62.00-78.00) | **<0.001** |
| Length-of-stay, median (IQR) | 3.00 (2.00-6.00) | 3.00 (2.00-6.00) | 4.00 (2.00-7.00) | 4.00 (2.00-7.00) | **<0.001** |
| Sex Female, N (%) | 557595 (50.41) | 538635 (50.47) | 9795 (46.88) | 9165 (51.03) | **<0.001** |
| **Ethnicity** | | | | | **<0.001** |
| White | 735330 (66.48) | 707550 (66.30) | 15005 (71.81) | 12775 (71.13) | |
| Black | 183090 (16.55) | 177790 (16.66) | 2880 (13.78) | 2420 (13.47) | |
| Hispanic | 84950 (7.68) | 82700 (7.75) | 1240 (5.93) | 1010 (5.62) | |
| Asian or Pacific Islander | 31635 (2.86) | 30500 (2.86) | 530 (2.54) | 605 (3.37) | |
| Native American | 4700 (0.42) | 4570 (0.43) | 60 (0.29) | 70 (0.39) | |
| Other | 27460 (2.48) | 26535 (2.49) | 410 (1.96) | 515 (2.87) | |
| **ELIXHAUSER CO-MORBIDITIES**, N (%) | | | | | |
| Congestive Heart Failure | 172170 (15.57) | 166950 (15.64) | 3225 (15.43) | 1995 (11.11) | **<0.001** |
| Valvular Disease | 110540 (9.99) | 106640 (9.99) | 2300 (11.01) | 1600 (8.91) | **0.008** |
| Pulmonary Circulation Disease | 8460 (0.76) | 6710 (0.63) | 555 (2.66) | 1195 (6.65) | **<0.001** |
| Peripheral Vascular Disease | 112065 (10.13) | 107955 (10.12) | 2470 (11.82) | 1640 (9.13) | **<0.001** |
| Paralysis | 112895 (10.21) | 108920 (10.21) | 2260 (10.82) | 1715 (9.55) | 0.178 |
| Other Neurological Disorders | 6620 (0.60) | 6275 (0.59) | 210 (1.01) | 135 (0.75) | **0.001** |

| | | | | | |
|---|---|---|---|---|---|
| Chronic Pulmonary Disease | 174180 (15.75) | 165690 (15.53) | 4795 (22.95) | 3695 (20.57) | **<0.001** |
| Diabetes (without chronic complications) | 210220 (19.01) | 203765 (19.09) | 3520 (16.85) | 2935 (16.34) | **<0.001** |
| Diabetes (with chronic complications) | 214400 (19.38) | 209135 (19.60) | 3095 (14.81) | 2170 (12.08) | **<0.001** |
| Hypothyroidism | 159160 (14.39) | 153810 (14.41) | 2950 (14.12) | 2400 (13.36) | 0.193 |
| Renal Failure | 181950 (16.45) | 175960 (16.49) | 3670 (17.56) | 2320 (12.92) | **<0.001** |
| Liver Disease | 18310 (1.66) | 17275 (1.62) | 550 (2.63) | 485 (2.70) | **<0.001** |
| Peptic Ulcer Disease | 7695 (0.70) | 7400 (0.69) | 175 (0.84) | 120 (0.67) | 0.527 |
| Acquired Immune Deficiency Syndrome | 2395 (0.22) | 2280 (0.21) | 100 (0.48) | 15 (0.08) | **<0.001** |
| Lymphoma | 5315 (0.48) | 4915 (0.46) | 220 (1.05) | 180 (1.00) | **<0.001** |
| Rheumatoid Arthritis / Collagen Vascular Disease | 30150 (2.73) | 29240 (2.74) | 520 (2.49) | 390 (2.17) | 0.075 |
| Coagulopathy | 41405 (3.74) | 37105 (3.48) | 1560 (7.47) | 2740 (15.26) | **<0.001** |
| Obesity | 145465 (13.15) | 142575 (13.36) | 1775 (8.49) | 1115 (6.21) | **<0.001** |
| Weight loss | 44030 (3.98) | 39685 (3.72) | 1795 (8.59) | 2550 (14.20) | **<0.001** |
| Fluid and electrolyte disorders | 246680 (22.30) | 235750 (22.09) | 5200 (24.89) | 5730 (31.90) | **<0.001** |
| Anaemia (chronic blood loss) | 4025 (0.36) | 3590 (0.34) | 240 (1.15) | 195 (1.09) | **<0.001** |
| Anaemia (deficiency) | 133005 (12.03) | 123720 (11.59) | 4375 (20.94) | 4910 (27.34) | **<0.001** |
| Alcohol abuse | 49375 (4.46) | 48080 (4.51) | 800 (3.83) | 495 (2.76) | **<0.001** |
| Drug abuse | 28985 (2.62) | 28365 (2.66) | 355 (1.70) | 265 (1.48) | **<0.001** |
| Psychoses | 26255 (2.37) | 25490 (2.39) | 440 (2.11) | 325 (1.81) | **0.041** |
| Depression | 124635 (11.27) | 120280 (11.27) | 2330 (11.15) | 2025 (11.28) | 0.971 |
| Hypertension | 946140 (85.54) | 916430 (85.87) | 16975 (81.24) | 12735 (70.91) | **<0.001** |

| **OTHER CO-MORBIDITIES**, N (%) | | | | | |
|---|---|---|---|---|---|
| Sepsis | 16400 (1.48) | 15275 (1.43) | 480 (2.30) | 645 (3.59) | **<0.001** |
| Liver Disease | 25520 (2.31) | 24165 (2.26) | 705 (3.37) | 650 (3.62) | **<0.001** |
| Anaemia | 151005 (13.65) | 138965 (13.02) | 5505 (26.35) | 6535 (36.39) | **<0.001** |
| Dyslipidaemia | 640010 (57.86) | 621485 (58.24) | 10800 (51.69) | 7725 (43.01) | **<0.001** |
| Dementia | 131650 (11.90) | 128305 (12.02) | 2365 (11.32) | 980 (5.46) | **<0.001** |
| Smoking | 203440 (18.39) | 197625 (18.52) | 3200 (15.31) | 2615 (14.56) | **<0.001** |
| Parkinson Disease | 15795 (1.43) | 15335 (1.44) | 315 (1.51) | 145 (0.81) | **0.006** |
| Transient Ischaemic Attack | 8070 (0.73) | 7835 (0.73) | 135 (0.65) | 100 (0.56) | 0.394 |
| Rheumatic heart Disease | 32355 (2.93) | 31205 (2.92) | 735 (3.52) | 415 (2.31) | **0.006** |
| Coronary Heart Disease | 314870 (28.47) | 303765 (28.46) | 6265 (29.98) | 4840 (26.95) | **0.013** |
| Heart Failure | 195835 (17.71) | 189915 (17.80) | 3600 (17.23) | 2320 (12.92) | **<0.001** |
| Pulmonary Embolism | 7000 (0.63) | 5390 (0.51) | 510 (2.44) | 1100 (6.12) | **<0.001** |
| Congenital Heart Disease | 33495 (3.03) | 32350 (3.03) | 545 (2.61) | 600 (3.34) | 0.169 |
| Pericarditis | 105 (0.01) | 90 (0.01) | <11* | <11* | **0.018** |
| Infectious Endocarditis | 2310 (0.21) | 2050 (0.19) | 70 (0.34) | 190 (1.06) | **<0.001** |
| Atrial Fibrillation | 282175 (25.51) | 272015 (25.49) | 6315 (30.22) | 3845 (21.41) | **<0.001** |
| Arrhythmias (other than Atrial Fibrillation) | 91590 (8.28) | 88440 (8.29) | 1925 (9.21) | 1225 (6.82) | **0.001** |
| Peripheral Vascular Disease | 117895 (10.66) | 113575 (10.64) | 2575 (12.32) | 1745 (9.72) | **<0.001** |
| Deep Venous Thrombosis | 14845 (1.34) | 12395 (1.16) | 840 (4.02) | 1610 (8.96) | **<0.001** |
| Gastrointestinal Bleeding | 12675 (1.15) | 11600 (1.09) | 560 (2.68) | 515 (2.87) | **<0.001** |

| | | | | | |
|---|---|---|---|---|---|
| Pneumonia | 29905 (2.70) | 27575 (2.58) | 1040 (4.98) | 1290 (7.18) | **<0.001** |
| Chronic Lung Disease | 136505 (12.34) | 129015 (12.09) | 4220 (20.20) | 3270 (18.21) | **<0.001** |
| Chronic Obstructive Pulmonary Disease | 126505 (11.44) | 119750 (11.22) | 3810 (18.23) | 2945 (16.40) | **<0.001** |
| Ventricular Septal Defect | 335 (0.03) | 320 (0.03) | <11* | <11* | 0.802 |
| Atrial Septal Defect | 31295 (2.83) | 30205 (2.83) | 510 (2.44) | 580 (3.23) | 0.122 |
| Shock | 5070 (0.46) | 4830 (0.45) | 95 (0.45) | 145 (0.81) | **0.007** |
| Family history of cerebrovascular disease | 42725 (3.86) | 41775 (3.91) | 585 (2.80) | 365 (2.03) | **<0.001** |
| Family history of heart disease | 64000 (5.79) | 62450 (5.85) | 840 (4.02) | 710 (3.95) | **<0.001** |
| Previous cerebrovascular disease | 172600 (15.61) | 167510 (15.70) | 2920 (13.97) | 2170 (12.08) | **<0.001** |
| **PROCEDURES**, N (%) | | | | | |
| Thrombectomy | 34420 (3.11) | 33090 (3.10) | 670 (3.21) | 660 (3.67) | 0.139 |
| Thrombolysis | 103600 (9.37) | 101035 (9.47) | 1730 (8.28) | 835 (4.65) | **<0.001** |
| **OUTCOMES**, N (%) | | | | | |
| In-hospital mortality | 43545 (3.94) | 40545 (3.80) | 1230 (5.89) | 1770 (9.86) | **<0.001** |
| Length-of-stay >4 days | 380605 (34.41) | 363430 (34.05) | 8680 (41.54) | 8495 (47.30) | **<0.001** |
| Routine Discharge | 394105 (35.99) | 384490 (36.39) | 5600 (26.94) | 4015 (22.52) | **<0.001** |
| **OTHER CHARACTERISTICS**, N (%) | | | | | |
| **Year of admission** | | | | | 0.755 |
| 2015 | 112000 (10.13) | 107955 (10.12) | 2115 (10.12) | 1930 (10.75) | |
| 2016 | 488005 (44.12) | 471070 (44.14) | 9135 (43.72) | 7800 (43.43) | |
| 2017 | 506040 (45.75) | 488165 (45.74) | 9645 (46.16) | 8230 (45.82) | |
| **Hospital bedsize** | | | | | **<0.001** |
| Small | 174745 (15.80) | 169030 (15.84) | 3205 (15.34) | 2510 (13.98) | |

| | | | | | |
|---|---|---|---|---|---|
| Medium | 324895 (29.37) | 314100 (29.43) | 5955 (28.50) | 4840 (26.95) | |
| Large | 606405 (54.83) | 584060 (54.73) | 11735 (56.16) | 10610 (59.08) | |
| **Location/teaching status of hospital** | | | | | **<0.001** |
| Rural | 82535 (7.46) | 80035 (7.50) | 1505 (7.20) | 995 (5.54) | |
| Urban non-teaching | 266440 (24.09) | 258170 (24.19) | 4625 (22.13) | 3645 (20.30) | |
| Urban teaching | 757070 (68.45) | 728985 (68.31) | 14765 (70.66) | 13320 (74.16) | |
| **Region of hospital** | | | | | **<0.001** |
| Northeast | 198830 (17.98) | 190840 (17.88) | 4235 (20.27) | 3755 (20.91) | |
| Midwest | 238555 (21.57) | 229900 (21.54) | 4660 (22.30) | 3995 (22.24) | |
| South | 461280 (41.71) | 446670 (41.85) | 7945 (38.02) | 6665 (37.11) | |
| West | 207380 (18.75) | 199780 (18.72) | 4055 (19.41) | 3545 (19.74) | |
| **All Patient Refined DRG: Severity of Illness Subclass** | | | | | **<0.001** |
| Minor loss of function | 89400 (8.08) | 88465 (8.29) | 935 (4.47) | <11* | |
| Moderate loss of function | 541350 (48.94) | 529495 (49.62) | 8025 (38.41) | 3830 (21.33) | |
| Major loss of function | 365580 (33.05) | 346570 (32.48) | 8955 (42.86) | 10055 (55.99) | |
| Extreme loss of function | 109715 (9.92) | 102660 (9.62) | 2980 (14.26) | 4075 (22.69) | |
| **Disposition of the patient at discharge** | | | | | **<0.001** |
| Routine Discharge | 394105 (35.63) | 384490 (36.03) | 5600 (26.80) | 4015 (22.36) | |
| Transfer to short-term hospital | 31995 (2.89) | 30845 (2.89) | 645 (3.09) | 505 (2.81) | |
| Transfer to other facility: Includes Skilled Nursing Facility (SNF), | 468640 (42.37) | 451890 (42.34) | 9505 (45.49) | 7245 (40.34) | |

| | | | | |
|---|---|---|---|---|
| Intermediate Care Facility (ICF), Another Type of Facility | | | | |
| Home Health Care | 156825 (14.18) | 148725 (13.94) | 3805 (18.21) | 4295 (23.91) |
| Against Medical Advice | 10685 (0.97) | 10480 (0.98) | 90 (0.43) | 115 (0.64) |
| Died | 43545 (3.94) | 40545 (3.80) | 1230 (5.89) | 1770 (9.86) |
| Discharged alive, destination unknown | 250 (0.02) | 215 (0.02) | 20 (0.10) | 15 (0.08) |

**Supplemental Table IV**. Patient characteristics on admission, stratified by the most prevalent 5 types of cancer: respiratory cancers, prostate cancer, breast cancer, pancreatic cancer or colorectal cancer.

| | **Respiratory Cancers** | **Prostate Cancer** | **Breast Cancer** | **Pancreatic Cancer** | **Colorectal Cancer** |
|---|---|---|---|---|---|
| N | 9490 | 4960 | 3375 | 2640 | 2490 |
| Age, median (IQR) | 71.00 (63.00-79.00) | 79.00 (71.00-85.00) | 74.00 (67.00-83.00) | 71.00 (63.00-77.00) | 74.00 (65.00-82.00) |
| Length-of-stay, median (IQR) | 4.00 (3.00-7.00) | 3.00 (2.00-6.00) | 4.00 (2.00-6.00) | 4.00 (3.00-7.00) | 4.00 (2.00-8.00) |
| Sex<br>Female, N (%) | 4855 (51.16) | 0 (0.00) | 3355 (99.41) | 1320 (50.00) | 1290 (51.81) |
| **Ethnicity** | | | | | |
| White | 7050 (74.29) | 3265 (65.83) | 2375 (70.37) | 1780 (67.42) | 1615 (64.86) |
| Black | 1155 (12.17) | 935 (18.85) | 505 (14.96) | 435 (16.48) | 415 (16.67) |
| Hispanic | 445 (4.69) | 325 (6.55) | 205 (6.07) | 170 (6.44) | 215 (8.63) |
| Asian or Pacific Islander | 340 (3.58) | 100 (2.02) | 65 (1.93) | 85 (3.22) | 85 (3.41) |
| Native American | 15 (0.16) | 15 (0.30) | <11* | <11* | 20 (0.80) |
| Other | 220 (2.32) | 95 (1.92) | 70 (2.07) | 115 (4.36) | 35 (1.41) |
| **ELIXHAUSER CO-MORBIDITIES**, N (%) | | | | | |
| Congestive Heart Failure | 1215 (12.80) | 765 (15.42) | 480 (14.22) | 270 (10.23) | 460 (18.47) |
| Valvular Disease | 880 (9.27) | 590 (11.90) | 350 (10.37) | 260 (9.85) | 265 (10.64) |
| Pulmonary Circulation Disease | 620 (6.53) | 50 (1.01) | 50 (1.48) | 335 (12.69) | 110 (4.42) |
| Peripheral Vascular Disease | 1255 (13.22) | 545 (10.99) | 340 (10.07) | 195 (7.39) | 230 (9.24) |
| Paralysis | 1105 (11.64) | 500 (10.08) | 285 (8.44) | 225 (8.52) | 275 (11.04) |
| Other Neurological Disorders | 70 (0.74) | 45 (0.91) | 20 (0.59) | 15 (0.57) | 45 (1.81) |

| | | | | | |
|---|---|---|---|---|---|
| Chronic Pulmonary Disease | 3745 (39.46) | 680 (13.71) | 580 (17.19) | 325 (12.31) | 390 (15.66) |
| Diabetes (without chronic complications) | 1370 (14.44) | 835 (16.83) | 580 (17.19) | 700 (26.52) | 440 (17.67) |
| Diabetes (with chronic complications) | 890 (9.38) | 740 (14.92) | 445 (13.19) | 360 (13.64) | 400 (16.06) |
| Hypothyroidism | 1090 (11.49) | 415 (8.37) | 675 (20.00) | 325 (12.31) | 355 (14.26) |
| Renal Failure | 1180 (12.43) | 1065 (21.47) | 435 (12.89) | 300 (11.36) | 420 (16.87) |
| Liver Disease | 175 (1.84) | 85 (1.71) | 40 (1.19) | 80 (3.03) | 85 (3.41) |
| Peptic Ulcer Disease | 35 (0.37) | 35 (0.71) | 70 (2.07) | 5 (0.19) | 25 (1.00) |
| Acquired Immune Deficiency Syndrome | 25 (0.26) | 15 (0.30) | <11* | <11* | <11* |
| Lymphoma | 85 (0.90) | 15 (0.30) | 25 (0.74) | 20 (0.76) | 30 (1.20) |
| Rheumatoid Arthritis / Collagen Vascular Disease | 310 (3.27) | 85 (1.71) | 115 (3.41) | 50 (1.89) | 70 (2.81) |
| Coagulopathy | 1170 (12.33) | 340 (6.85) | 210 (6.22) | 500 (18.94) | 230 (9.24) |
| Obesity | 415 (4.37) | 395 (7.96) | 390 (11.56) | 205 (7.77) | 215 (8.63) |
| Weight loss | 1270 (13.38) | 290 (5.85) | 225 (6.67) | 420 (15.91) | 300 (12.05) |
| Fluid and electrolyte disorders | 2740 (28.87) | 1150 (23.19) | 910 (26.96) | 885 (33.52) | 750 (30.12) |
| Anaemia (chronic blood loss) | 60 (0.63) | 20 (0.40) | <11* | <11* | 160 (6.43) |
| Anaemia (deficiency) | 2400 (25.29) | 1070 (21.57) | 590 (17.48) | 795 (30.11) | 870 (34.94) |
| Alcohol abuse | 365 (3.85) | 180 (3.63) | 65 (1.93) | 80 (3.03) | 100 (4.02) |
| Drug abuse | 165 (1.74) | 70 (1.41) | 30 (0.89) | 20 (0.76) | 25 (1.00) |
| Psychoses | 205 (2.16) | 85 (1.71) | 105 (3.11) | 45 (1.70) | 35 (1.41) |
| Depression | 1070 (11.28) | 435 (8.77) | 475 (14.07) | 320 (12.12) | 255 (10.24) |
| Hypertension | 6750 (71.13) | 4230 (85.28) | 2720 (80.59) | 1870 (70.83) | 1980 (79.52) |

| **OTHER CO-MORBIDITIES**, N (%) | | | | | |
|---|---|---|---|---|---|
| Sepsis | 280 (2.95) | 100 (2.02) | 75 (2.22) | 130 (4.92) | 125 (5.02) |
| Liver Disease | 215 (2.27) | 100 (2.02) | 55 (1.63) | 115 (4.36) | 100 (4.02) |
| Anaemia | 3080 (32.46) | 1215 (24.50) | 790 (23.41) | 975 (36.93) | 1205 (48.39) |
| Dyslipidaemia | 4355 (45.89) | 2725 (54.94) | 1580 (46.81) | 1130 (42.80) | 1160 (46.59) |
| Dementia | 550 (5.80) | 780 (15.73) | 305 (9.04) | 95 (3.60) | 240 (9.64) |
| Smoking | 2145 (22.60) | 500 (10.08) | 430 (12.74) | 335 (12.69) | 280 (11.24) |
| Parkinson Disease | 70 (0.74) | 125 (2.52) | 35 (1.04) | <11* | 15 (0.60) |
| Transient Ischaemic Attack | 30 (0.32) | 60 (1.21) | 25 (0.74) | <11* | <11* |
| Rheumatic Heart Disease | 270 (2.85) | 175 (3.53) | 140 (4.15) | 60 (2.27) | 75 (3.01) |
| Coronary Heart Disease | 2960 (31.19) | 1715 (34.58) | 700 (20.74) | 780 (29.55) | 745 (29.92) |
| Heart Failure | 1355 (14.28) | 905 (18.25) | 535 (15.85) | 305 (11.55) | 530 (21.29) |
| Pulmonary Embolism | 575 (6.06) | 45 (0.91) | 40 (1.19) | 325 (12.31) | 100 (4.02) |
| Congenital Heart Disease | 330 (3.48) | 110 (2.22) | 60 (1.78) | 110 (4.17) | 30 (1.20) |
| Pericarditis | <11* | <11* | <11* | <11* | <11* |
| Infectious Endocarditis | 80 (0.84) | <11* | 25 (0.74) | 40 (1.52) | <11* |
| Atrial Fibrillation | 2330 (24.55) | 1595 (32.16) | 975 (28.89) | 535 (20.27) | 710 (28.51) |
| Arrhythmias (other than Atrial Fibrillation) | 730 (7.69) | 545 (10.99) | 285 (8.44) | 165 (6.25) | 190 (7.63) |
| Peripheral Vascular Disease | 1270 (13.38) | 565 (11.39) | 355 (10.52) | 205 (7.77) | 230 (9.24) |
| Deep Venous Thrombosis | 795 (8.38) | 90 (1.81) | 75 (2.22) | 405 (15.34) | 110 (4.42) |
| Gastrointestinal Bleeding | 145 (1.53) | 45 (0.91) | 30 (0.89) | 65 (2.46) | 245 (9.84) |

| | | | | | |
|---|---|---|---|---|---|
| Pneumonia | 1015 (10.70) | 135 (2.72) | 165 (4.89) | 125 (4.73) | 105 (4.22) |
| Chronic Lung Disease | 3625 (38.20) | 590 (11.90) | 465 (13.78) | 265 (10.04) | 310 (12.45) |
| Chronic Obstructive Pulmonary Disease | 3310 (34.88) | 505 (10.18) | 440 (13.04) | 230 (8.71) | 265 (10.64) |
| Ventricular Septal Defect | <11* | <11* | <11* | <11* | <11* |
| Atrial Septal Defect | 315 (3.32) | 110 (2.22) | 60 (1.78) | 105 (3.98) | 30 (1.20) |
| Shock | 100 (1.05) | 15 (0.30) | 15 (0.44) | 20 (0.76) | 15 (0.60) |
| Family history of cerebrovascular disease | 235 (2.48) | 120 (2.42) | 90 (2.67) | 50 (1.89) | 95 (3.82) |
| Family history of heart disease | 385 (4.06) | 190 (3.83) | 140 (4.15) | 85 (3.22) | 90 (3.61) |
| Previous cerebrovascular disease | 1190 (12.54) | 730 (14.72) | 455 (13.48) | 300 (11.36) | 295 (11.85) |
| **PROCEDURES**, N (%) | | | | | |
| Thrombectomy | 405 (4.27) | 130 (2.62) | 80 (2.37) | 120 (4.55) | 90 (3.61) |
| Thrombolysis | 445 (4.69) | 440 (8.87) | 320 (9.48) | 135 (5.11) | 145 (5.82) |
| **OUTCOMES**, N (%) | | | | | |
| In-hospital mortality | 965 (10.17) | 185 (3.73) | 190 (5.63) | 280 (10.61) | 155 (6.22) |
| Length-of-stay >4 days | 4470 (47.10) | 1850 (37.30) | 1280 (37.93) | 1295 (49.05) | 1170 (46.99) |
| Routine Discharge | 2010 (21.31) | 1505 (30.47) | 995 (29.57) | 475 (18.06) | 635 (25.76) |
| **OTHER CHARACTERISTICS**, N (%) | | | | | |
| **Year of admission** | | | | | |
| 2015 | 935 (9.85) | 530 (10.69) | 345 (10.22) | 310 (11.74) | 280 (11.24) |
| 2016 | 4265 (44.94) | 2180 (43.95) | 1275 (37.78) | 1145 (43.37) | 1035 (41.57) |
| 2017 | 4290 (45.21) | 2250 (45.36) | 1755 (52.00) | 1185 (44.89) | 1175 (47.19) |
| **Hospital bedsize** | | | | | |
| Small | 1225 (12.91) | 820 (16.53) | 550 (16.30) | 395 (14.96) | 410 (16.47) |

| | | | | | |
|---|---|---|---|---|---|
| Medium | 2740 (28.87) | 1445 (29.13) | 1060 (31.41) | 695 (26.33) | 695 (27.91) |
| Large | 5525 (58.22) | 2695 (54.33) | 1765 (52.30) | 1550 (58.71) | 1385 (55.62) |
| **Location/teaching status of hospital** | | | | | |
| Rural | 540 (5.69) | 335 (6.75) | 325 (9.63) | 150 (5.68) | 175 (7.03) |
| Urban non-teaching | 2130 (22.44) | 1175 (23.69) | 740 (21.93) | 460 (17.42) | 535 (21.49) |
| Urban teaching | 6820 (71.87) | 3450 (69.56) | 2310 (68.44) | 2030 (76.89) | 1780 (71.49) |
| **Region of hospital** | | | | | |
| Northeast | 1890 (19.92) | 1070 (21.57) | 680 (20.15) | 625 (23.67) | 405 (16.27) |
| Midwest | 2180 (22.97) | 1095 (22.08) | 780 (23.11) | 475 (17.99) | 575 (23.09) |
| South | 3765 (39.67) | 1755 (35.38) | 1285 (38.07) | 1010 (38.26) | 1080 (43.37) |
| West | 1655 (17.44) | 1040 (20.97) | 630 (18.67) | 530 (20.08) | 430 (17.27) |
| **All Patient Refined DRG: Severity of Illness Subclass** | | | | | |
| Minor loss of function | 130 (1.37) | 280 (5.65) | 165 (4.89) | 20 (0.76) | 50 (2.01) |
| Moderate loss of function | 2120 (22.34) | 2130 (42.94) | 1415 (41.93) | 590 (22.35) | 780 (31.33) |
| Major loss of function | 5015 (52.85) | 2010 (40.52) | 1410 (41.78) | 1450 (54.92) | 1170 (46.99) |
| Extreme loss of function | 2225 (23.45) | 540 (10.89) | 385 (11.41) | 580 (21.97) | 490 (19.68) |
| **Disposition of the patient at discharge** | | | | | |
| Routine Discharge | 2010 (21.18) | 1505 (30.34) | 995 (29.48) | 475 (17.99) | 635 (25.50) |
| Transfer to short-term hospital | 285 (3.00) | 180 (3.63) | 40 (1.19) | 70 (2.65) | 45 (1.81) |
| Transfer to other facility: Includes Skilled Nursing Facility (SNF), | 3950 (41.62) | 2275 (45.87) | 1440 (42.67) | 985 (37.31) | 1130 (45.38) |

| | | | | | |
|---|---|---|---|---|---|
| Intermediate Care Facility (ICF), Another Type of Facility | | | | | |
| Home Health Care | 2220 (23.39) | 795 (16.03) | 700 (20.74) | 820 (31.06) | 500 (20.08) |
| Against Medical Advice | 50 (0.53) | 15 (0.30) | <11* | <11* | 15 (0.60) |
| Died | 965 (10.17) | 185 (3.73) | 190 (5.63) | 280 (10.61) | 155 (6.22) |
| Discharged alive, destination unknown | <11* | <11* | <11* | <11* | <11* |

IQR – inter-quartile range; ***** cell sizes $\leq 10$ were not reported in order to avoid patient re-identification, according to the HCUP guidelines[21];

**Supplemental Table V.** Results of multivariable logistic regressions evaluating the associations between non-metastatic and metastatic cancer and the odds of receiving thrombolysis or thrombectomy for acute ischaemic stroke.

| | **Intravenous Thrombolysis** | **Endovascular Thrombectomy** |
|---|---|---|
| | **OR (95% CI)** | **OR (95% CI)** |
| No cancer | Reference | Reference |
| Non-metastatic cancer | **0.77 (0.66-0.90)** | **0.80 (0.63-0.9994)** |
| Metastatic cancer | **0.39 (0.32-0.47)** | 0.87 (0.69-1.09) |

Models adjusted for age, sex, ethnicity, Elixhauser co-morbidities (congestive heart failure, valvular disease, pulmonary circulatory disease, peripheral vascular disease, paralysis, other neurological disorders, chronic pulmonary disease, diabetes, hypothyroidism, renal failure, liver disease, peptic ulcer disease, acquired immune deficiency syndrome, rheumatoid arthritis, coagulopathy, obesity, weight loss, fluid and electrolyte disorders, anaemia, alcohol abuse, drug abuse, psychosis, depression and hypertension), previous history of cancer, haematological cancers, other co-morbidities (dyslipidaemia, smoking, Parkinson disease, coronary heart disease, all-cause bleeding, pulmonary embolism, deep venous thrombosis, atrial fibrillation, arrhythmias other than atrial fibrillation, pneumonia (incl. aspiration), shock, previous cerebrovascular disease), hospital bedsize, location & teaching status and other revascularisation therapy.

Statistically significant differences ($P$ <0.01) highlighted in **bold**.
